# Characterization and contextualization of transgender women population and condom use in the HIV syndemic framework: Scoping review

**DOI:** 10.1101/2023.07.07.23292379

**Authors:** Jorge Eduardo Moncayo-Quevedo, María Del Mar Pérez-Arizabaleta, Alejandra Rocio Rodríguez-Ortiz, Lina María Villegas-Trujillo

## Abstract

**Objective:** To characterize and contextualize condom use in the transgender women (TW) population utilizing the HIV syndemic framework.

**Methods:** Studies reported condom-use frequency and syndemic factors associated with HIV risk in the TW population were searched in databases. We followed the PRISMA Extension for Scoping Reviews checklist.

**Results:** Social factors have a proven relationship with using condoms and HIV among TW. Syndemic factors, and how some of them reinforce others, deserve a specific analysis to develop strategies to face HIV among TW.

**Conclusions:** Analyzing a syndemic perspective allows to generate specific health intervention and prevention policies to protect the TW.

## Introduction

Transgender women (TW) are a vulnerable population worldwide (Becasen et al., 2019; Yi et al., 2019). The term transgender is used most often to refer to people whose gender identity or expression differs from their birth sex; this document refers to people who were assigned male at birth, but identified as women (Baral et al., 2013; Poteat et al., 2017). As a result of the social marginalization, mental health challenges, violence, and drug abuse that TW suffer, they have higher rates of HIV than other populations (Chakrapani et al., 2017). In this context, the presence of psychosocial factors confers a higher risk of acquiring HIV, which relates to the concept of “syndemic” (Alvarado et al., 2020).

With syndemic approach, HIV can be understood as the synergic interaction of multiple psychosocial factors that amplify the negative impact of the disease (Singer et al., 2006; Tsai et al., 2017). In syndemic studies health conditions can be determined by social, economic, or political factors (Alvarado et al., 2020). This theory includes disease epidemics and social conditions contributing to a specific disease’s proliferation (Wilson et al., 2014). According to the study by Alvarado et al. (2020), the syndemic theory is a suitable approach to the effects of psychosocial conditions of HIV, given its complex nature. Moreover, Chakrapani et al. (2019) found in their study in India with 300 TW that syndemic conditions like depression, alcohol use, and violent victimization were significant mediators of the effect of transgender identity stigma on sexual health risks. Likewise, Alessi et al. (2020) used a syndemic approach to explore the processes that lead to heightened HIV risk in sexual and gender minorities from South Africa, finding that syndemic factors such as financial and housing instability, discrimination, food insecurity, and lack of social support among others, contribute to HIV risk. For this reason, this scoping review aims to characterize and contextualize condom use in the TW population under the HIV syndemic framework.

## Materials and Methods

### Protocol and registration

For this scoping review we followed the PRISMA Extension for Scoping Reviews (PRISMA-ScR) checklist (Tricco et al., 2018). The protocol for this review was registered in the Open Science Framework (OSF) database (Moncayo Quevedo et al., 2021).

### Eligibility criteria

Original studies, such as observational studies and surveys that reported condom-use frequency and syndemic factors associated with HIV risk in the TW population, were eligible for this scoping review. We did not restrain the search based on the time of publication. In the case of retrieving from the search multiple articles with the same sample, only the one that adhered the most to the subject of this review was included.

### Information sources and search

We performed a systematic search in multiple databases including Scopus, Web of Science (WoS), PubMed, OVID and Cochrane. Additionally, we used MeSH terms and non-controlled vocabulary that we considered fundamental to our objective: HIV or Human immunodeficiency virus, transgender women, condom and health, or unsafe sex. The search equation used was the following:

((TITLE-ABS-KEY (HIV) OR TITLE-ABS-KEY (human AND immunodeficiency AND virus*)))

AND

(TITLE-ABS-KEY (transgender AND women*))

AND

(TITLE-ABS-KEY (condom*))

AND

(LIMIT-TO (DOCTYPE, “ar”))

We did not add language or time restrictions to the searches.

### Selection of sources and data charting process

We included studies describing condom use frequency and syndemic factors associated with HIV risk in the TW population. To select the articles, first, two authors independently examined only the titles and abstracts of studies yielded from the searches to identify those that met the established criteria. After this initial selection, those articles were downloaded for full-text review. Once this screening ended, the same authors independently reviewed the full texts and decided whether they should be included or not in the scoping review based on the inclusion criteria. Finally, the data was extracted by the same researchers. Disagreements were resolved by reaching a consensus.

### Data items

The abstraction variables included: the author, year of the study, country, study design, total sample size, a sample size of TW, ethnicity, age, percentage of sex worker participants, condom use (%), and syndemic factors (%). Among the syndemic factors, we included: a history of incarceration, childhood sexual abuse, forced sex, violent victimization, frequent alcohol use, illicit drug use, financial instability, depression, unstable housing, completed high school, stigma, and discrimination.

### Synthesis of results and data analysis

A data extraction table was built in Microsoft Excel to organize the results. Also, a descriptive summary and a thematic analysis of the included articles were conducted to better display the scoping review results. The authors read the articles included and identified the topics in which the review could be presented. Based on those topics, the findings were organized for each study, highlighting the main results. Data from stigma and discrimination variables were shown in the narrative section of results but not in tables. The statistical analysis for categorical variables consisted of percentages, frequencies, and measures of central tendency.

## Results

### Selection of sources of evidence

The systematic search yielded 589 articles after removing duplicates. The abstracts and titles of these studies were screened, finding that only 63 articles were related to the topic we aimed to analyze. After reviewing the articles in full text and applying the inclusion/exclusion criteria, 16 were further excluded. The resulting 47 studies were included in this scoping review (Figure 1).

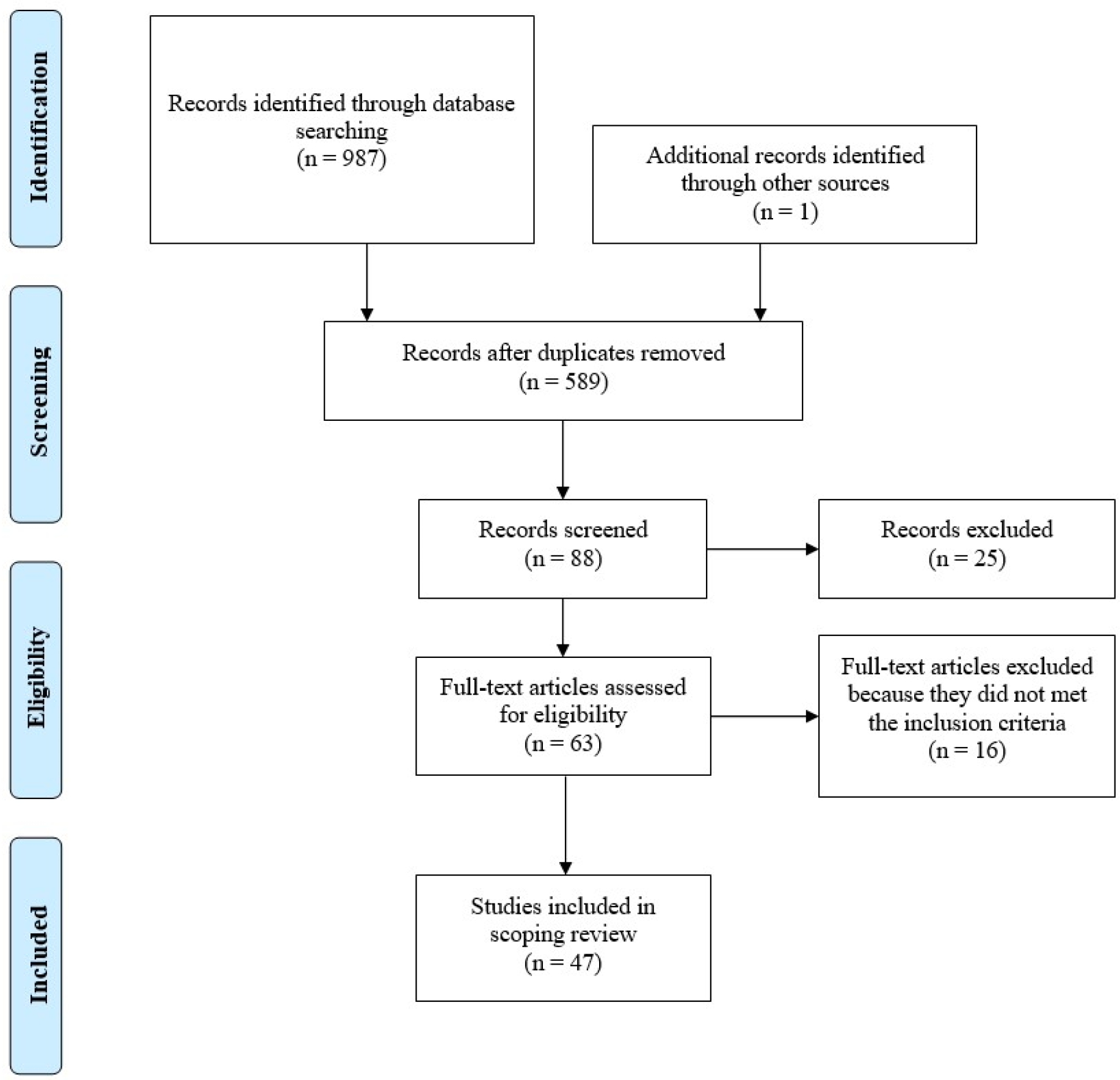

### Characteristics of sources of evidence

The main characteristics of the 47 selected papers are organized in supplementary table 1. To show a synthesized version of our results clearly and understandably, we present in Table 1 only a few of the selected studies that had data for all sociodemographic variables analyzed. The studies ranged from 2008 to 2020; The largest percentage of the selected papers were form the United States (23.4%) and had a cross-sectional design (57.4%). Moreover, most studies were focused on sex workers or had sex workers among the sample (61.7%) (Table 1).

**Table 1.**
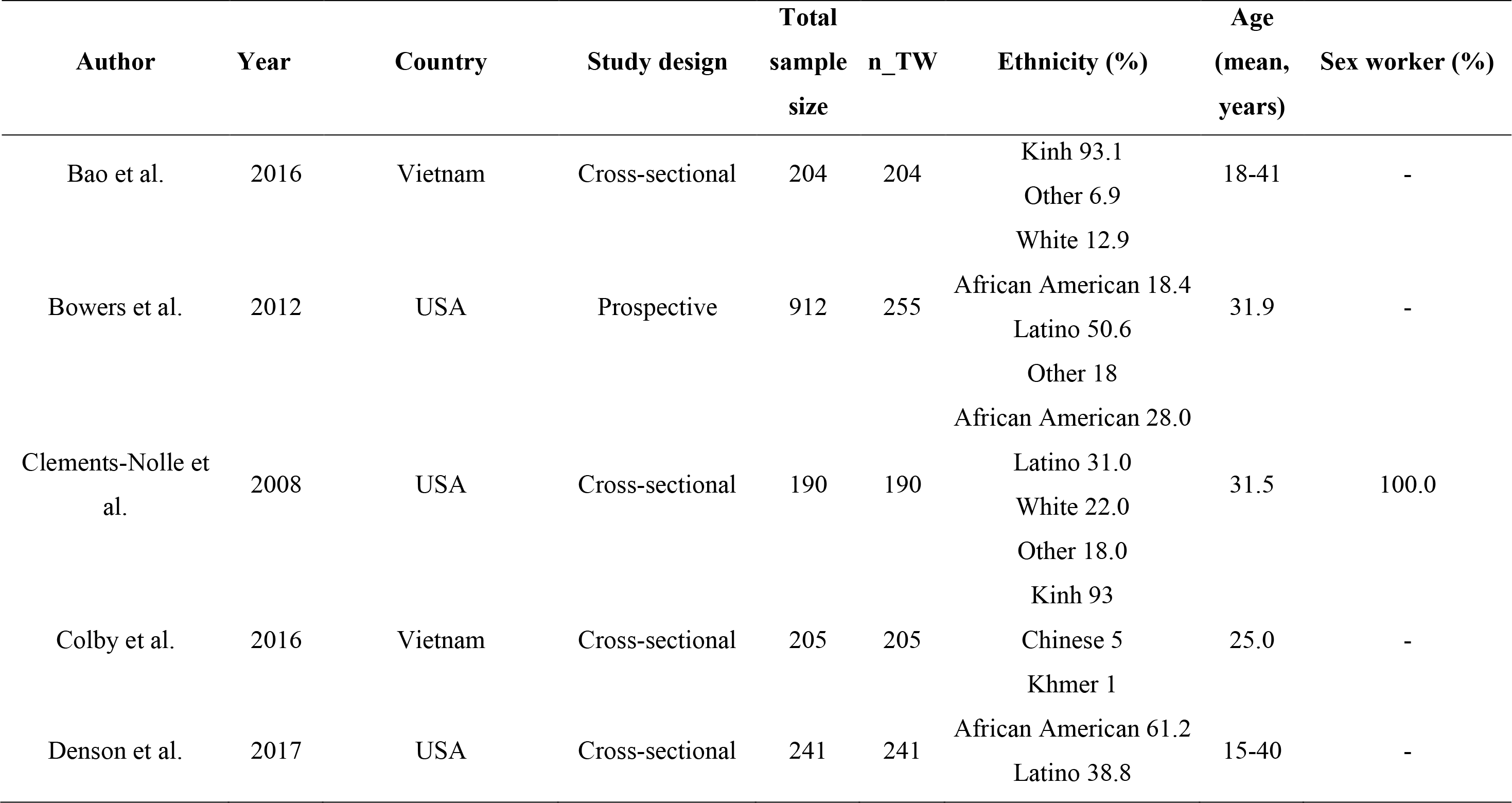
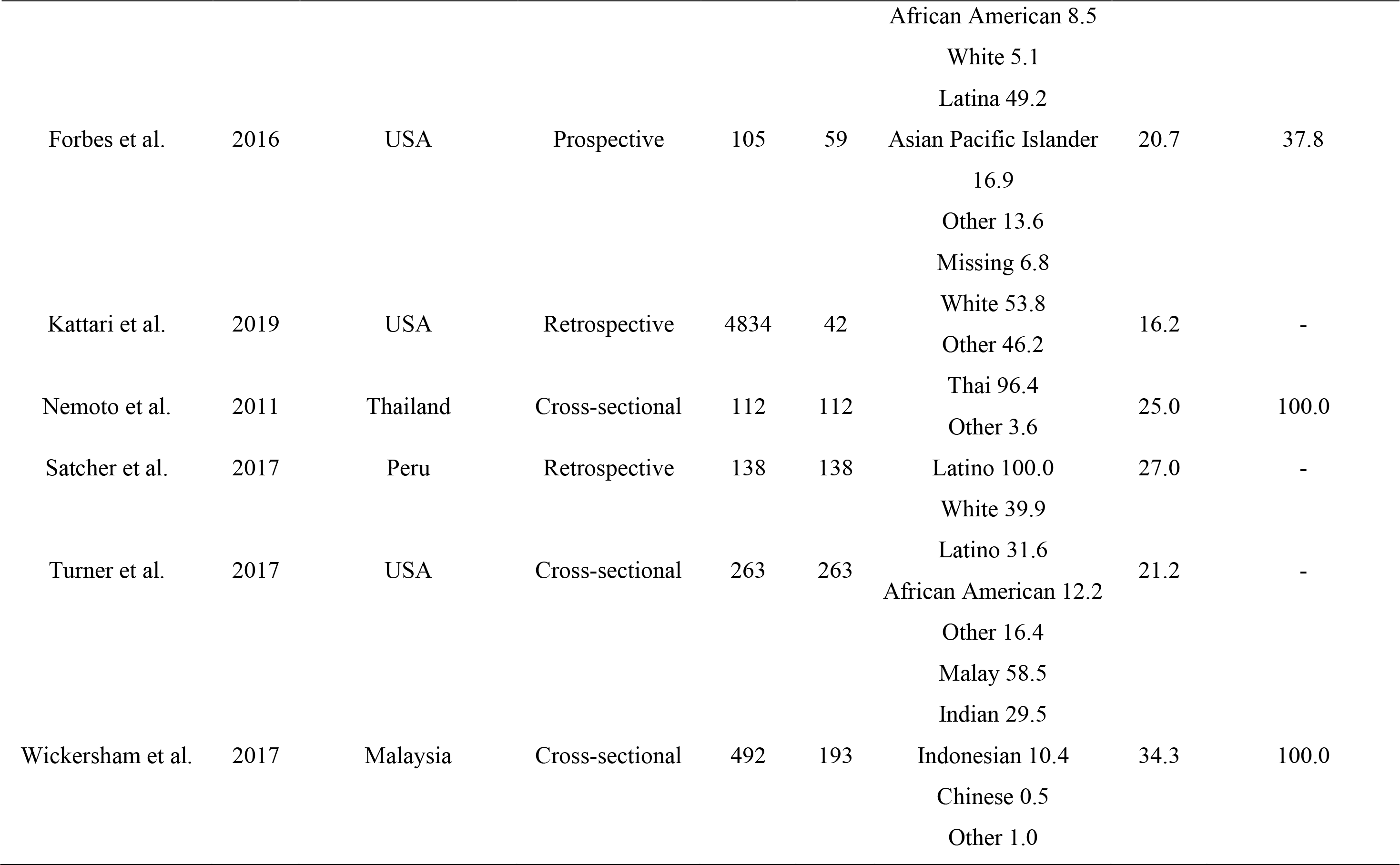
Characteristics of some of the included studies (Full table shown in supplementary material 1)

#### Condom use and syndemic factors

Results obtained from all variables analyzed in included studies regarding condom use and syndemic factors are presented in Supplementary Table 2 and Supplementary Table 3 respectively. In supplementary Table 2, only 26 studies are presented because only those reported condom use data. To present the results in a clear and synthesized version, Table 2 utilized only a few selected studies with data for most of the outcome variables analyzed.

**Table 2.**
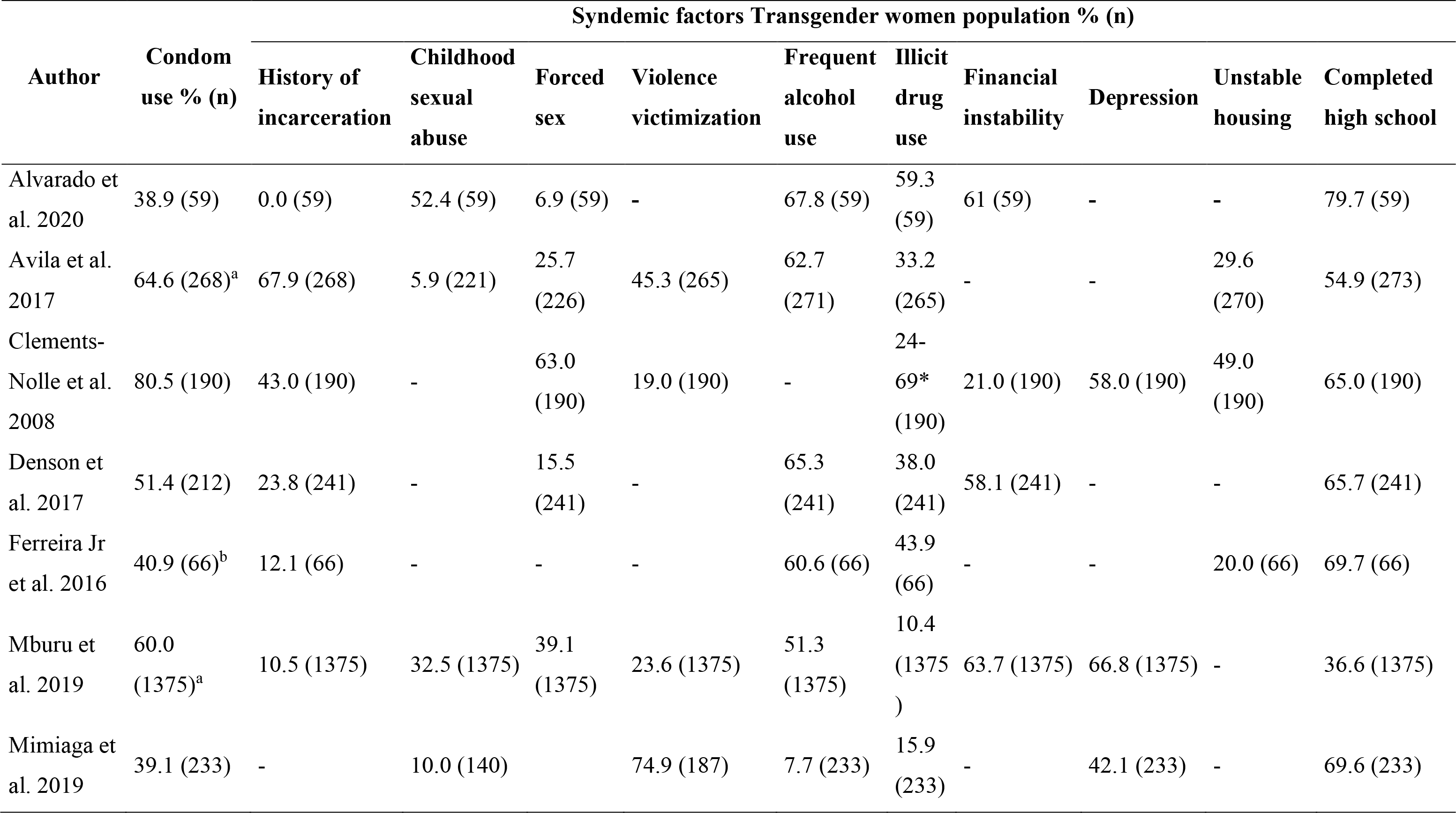
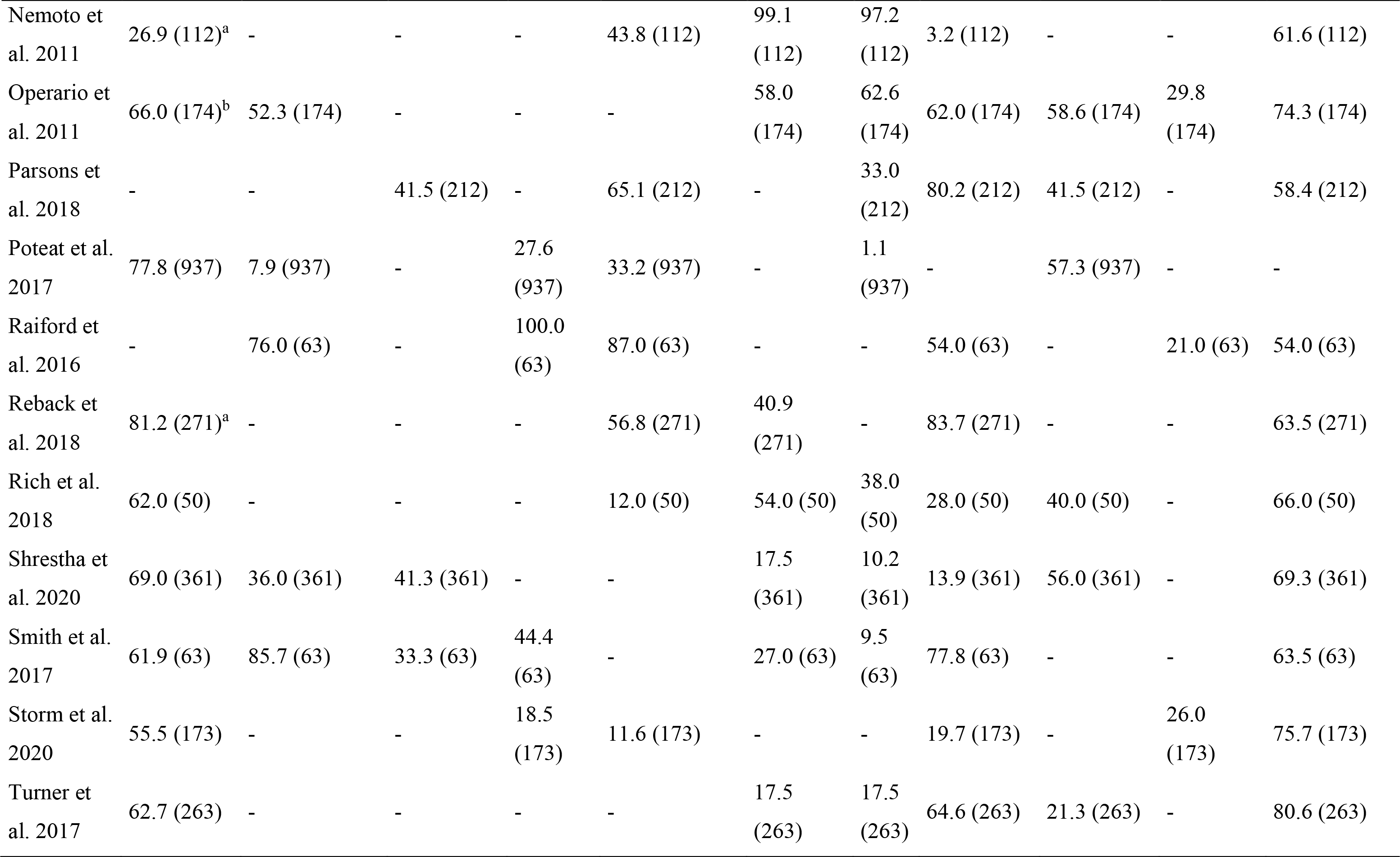
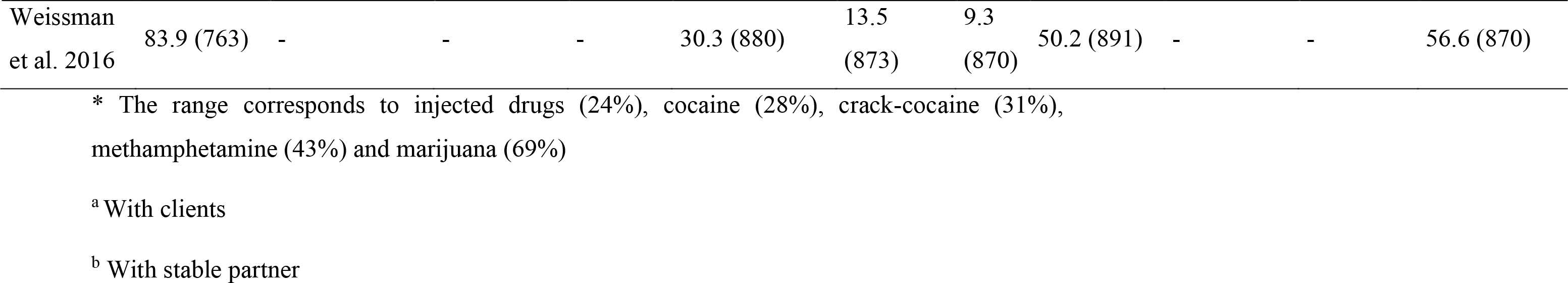
Syndemic factors and general condom use of some of the included studies (Full tables shown in supplementary material 2-3)

| Author | Condom use % (n) | Syndemic factors Transgender women population % (n) |  |  |  |  |  |  |  |  |  |
| --- | --- | --- | --- | --- | --- | --- | --- | --- | --- | --- | --- |
|  |  | History of incarceration | Childhood sexual abuse | Forced sex | Violence victimization | Frequent alcohol use | Illicit drug use | Financial instability | Depression | Unstable housing | Completed high school |
| Alvarado et al. 2020 | 38.9 (59) | 0.0 (59) | 52.4 (59) | 6.9 (59) | - | 67.8 (59) | 59.3 (59) | 61 (59) | - | - | 79.7 (59) |
| Avila et al. 2017 | 64.6 (268)<sup>a</sup> | 67.9 (268) | 5.9 (221) | 25.7 (226) | 45.3 (265) | 62.7 (271) | 33.2 (265) | - | - | 29.6 (270) | 54.9 (273) |
| Clements-Nolle et al. 2008 | 80.5 (190) | 43.0 (190) | - | 63.0 (190) | 19.0 (190) | - | 24-69* (190) | 21.0 (190) | 58.0 (190) | 49.0 (190) | 65.0 (190) |
| Denson et al. 2017 | 51.4 (212) | 23.8 (241) | - | 15.5 (241) | - | 65.3 (241) | 38.0 (241) | 58.1 (241) | - | - | 65.7 (241) |
| Ferreira Jr et al. 2016 | 40.9 (66)<sup>b</sup> | 12.1 (66) | - | - | - | 60.6 (66) | 43.9 (66) | - | - | 20.0 (66) | 69.7 (66) |
| Mburu et al. 2019 | 60.0 (1375)<sup>a</sup> | 10.5 (1375) | 32.5 (1375) | 39.1 (1375) | 23.6 (1375) | 51.3 (1375) | 10.4 (1375) | 63.7 (1375) | 66.8 (1375) | - | 36.6 (1375) |
| Mimiaga et al. 2019 | 39.1 (233) | - | 10.0 (140) | - | 74.9 (187) | 7.7 (233) | 15.9 (233) | - | 42.1 (233) | - | 69.6 (233) |
| Nemoto et al. 2011 | 26.9 (112)<sup>a</sup> | - | - | - | 43.8 (112) | 99.1 (112) | 97.2 (112) | 3.2 (112) | - | - | 61.6 (112) |
| Operario et al. 2011 | 66.0 (174)<sup>b</sup> | 52.3 (174) | - | - | - | 58.0 (174) | 62.6 (174) | 62.0 (174) | 58.6 (174) | 29.8 (174) | 74.3 (174) |
| Parsons et al. 2018 | - | - | 41.5 (212) | - | 65.1 (212) | - | 33.0 (212) | 80.2 (212) | 41.5 (212) | - | 58.4 (212) |
| Poteat et al. 2017 | 77.8 (937) | 7.9 (937) | - | 27.6 (937) | 33.2 (937) | - | 1.1 (937) | - | 57.3 (937) | - | - |
| Raiford et al. 2016 | - | 76.0 (63) | - | 100.0 (63) | 87.0 (63) | - | - | 54.0 (63) | - | 21.0 (63) | 54.0 (63) |
| Reback et al. 2018 | 81.2 (271)<sup>a</sup> | - | - | - | 56.8 (271) | 40.9 (271) | - | 83.7 (271) | - | - | 63.5 (271) |
| Rich et al. 2018 | 62.0 (50) | - | - | - | 12.0 (50) | 54.0 (50) | 38.0 (50) | 28.0 (50) | 40.0 (50) | - | 66.0 (50) |
| Shrestha et al. 2020 | 69.0 (361) | 36.0 (361) | 41.3 (361) | - | - | 17.5 (361) | 10.2 (361) | 13.9 (361) | 56.0 (361) | - | 69.3 (361) |
| Smith et al. 2017 | 61.9 (63) | 85.7 (63) | 33.3 (63) | 44.4 (63) | - | 27.0 (63) | 9.5 (63) | 77.8 (63) | - | - | 63.5 (63) |
| Storm et al. 2020 | 55.5 (173) | - | - | 18.5 (173) | 11.6 (173) | - | - | 19.7 (173) | - | 26.0 (173) | 75.7 (173) |
| Turner et al. 2017 | 62.7 (263) | - | - | - | - | 17.5 (263) | 17.5 (263) | 64.6 (263) | 21.3 (263) | - | 80.6 (263) |
| Weissman<br>et al. 2016 | 83.9 (763) | - | - | - | 30.3 (880) | 13.5<br>(873) | 9.3<br>(870) | 50.2 (891) | - | - | 56.6 (870) |

\* The range corresponds to injected drugs (24%), cocaine (28%), crack-cocaine (31%), methamphetamine (43%) and marijuana (69%)
<sup>a</sup> With clients
<sup>b</sup> With stable partner

### Synthesis of results

#### Condom use

##### General condom use

Not all studies showed results regarding condom use; only 26 reported any use, including the use with a stable partner, casual partner, and clients (Supplementary Table 2). Of those studies, 15 (57.7%) showed a general percentage of condom use self-reported by the participants. Regarding these studies that measure condom use in general, it was found that the highest condom use was observed in the study by Wickersham et al. (2017), which evaluated the prevalence of HIV among cisgender and transgender sex workers in Malaysia. According to the authors, the data associated with condom use show a significant difference between cis and trans women, with condom use greater in trans women (89%) than cis women (75.4%). On the other hand, Hearld et al. (2019) in the Dominican Republic examined the associations between alcohol consumption, high-risk sexual behaviors, and experiences of stigma among TW based on the assumption that the greater the consumption of alcohol, the greater the probability of risky behaviors. However, according to the result of this study, using condoms in anal intercourse in the last 30 days was 86.22%. Along these same lines, the study by Weissman et al. (2016) conducted with transgender people from Cambodia found that the general use of condoms during the last anal intercourse with a man was 83.9%. However, these results vary regarding soliciting or paying for sex. On the other hand, in the study by Clements-Nolle et al. (2008), carried out in the United States, a consistency of condom use of 80.5% was found in anal intercourse among transgender sex workers within the last six months.

We found the lowest percentages of condom use in the studies by Habarta et al. (2015), Khan et al. (2008), and Moriarty et al. (2019). In the research carried out by Habarta et al. (2015) in the United States, a percentage of condom use of 23.0% was observed in transgender people. Khan et al. (2008) reported a significantly lower condom use, of 15.0% in their study on HIV prevalence and sexually transmitted infections among Hijras in Pakistan. Moreover, the study by Moriarty et al. (2019), carried out in Peru, found a 10.1% of condom use in sexual relations in the last 30 days.

#### Syndemic factors

##### History of incarceration

Eleven studies informed percentage data of incarceration history self-reported by TW (Table 2). Denson et al. (2017) found that 24% of TW were in prison in the last 12 months; Operario et al. (2008) showed that 52% of the study’s population had been in prison at some point. The incarceration of transgender population is common: 27% were in prison in the last 12 months, and 58.7% were imprisoned at some point in their lives (Operario et al., 2008). The study by Ferreira et al (2016), showed that 12.1% of TW in Sao Paulo were in prison at least once.

Raiford et al. (2016) showed that 76% of participants had been in jail at least once, and 21% were incarcerated in the last three months. In the same way, Avila et al. (2017) asserted evidence that women who had once been arrested had a higher chance of having syphilis (OR 2.40) and HIV (1.42). The study by Wickersham et al. (2017) further explored incarceration, evidencing that 50.3% of TW were in jail at least once in their lives, and 26.9% went to prison mainly due to sex work (43.3%), using drugs (18.6%) and selling drugs (2.1%).

Regarding the use of condoms and incarceration, those women who had been in prison in the last six months presented a higher, inconsistent use of condoms (59%) than those in liberty (41%). At the same time, the practice of consistent condom use is higher among women in freedom (61%) than those in jail (39%), according to a study by Clements-Nolle et al. (2008). Mburu et al. (2019) showed that 10.5% of TW were arrested solely based on their gender identity expression. Furthermore, there is a difference between the time in prison or a temporary arrest, and according to the study by Shrestha et al. (2020), who showed that 36.6% of TW were in jail and 21.3% were in prison. On the other hand, Poteat et al. (2017) study evidenced that 7.9% of the population had an arrest record and 5.2% had been imprisoned.

##### Childhood sexual abuse (CSA)

Nine studies showed percentage data of childhood abuse self-reported by TW (Supplementary Table 2). Shrestha et al. (2020) reported that 41.3% of TW suffer from CSA (Shrestha et al., 2020). Likewise, Mburu et al. (2019) found that CSA was present in 32.5% of TW, and the percentage of recent drug use is lower in those not abused (43.4%) than those abused (56.6%) (Mburu et al., 2019).

Avila et al. (2017) reported that 5.9% of TW go through CSA, and those who had their first sexual intercourse before 13 y/o had a higher prevalence of syphilis (60%) and HIV (41.8%) than the TW with the first sexual intercourse after 13 y/o (43.3% and 28.1% respectively) (Avila et al., 2017).

##### Forced sex

Twelve studies showed percentage data of forced sex self-reported by TW (Supplementary Table 2). Avila et al. (2017) reported that 25.7% of the population had suffered from forced sex (Avila et al., 2017). Similar proportions were shown in Poteat’s study (27.6%) (Poteat et al., 2017) and in Mburu’s (39.1%) (Mburu et al., 2019). The study by Clements-Nolle et al. (2008) reported a high level of sexual violence; 63% of TW were forced to have sex or raped (Clements-Nolle et al., 2008).

Raiford et al. (2016) evidenced that all the TW were forced to have sex, and 68% of those events were recent (Raiford et al., 2016). Denson et al. (2017) present lower data, showing that in the past 12 months, by the time of the study, 15.5% of TW were forced to have sex.

##### Violence victimization

Twenty studies presented percentage data of violence victimization self-reported by TW (Supplementary Table 2). According to the study by Avila et al. (2017), the victims of physical violence (45.3% of the study’s population) had a higher prevalence of HIV (Avila et al., 2017). Clements-Nole’s study reflected that 19% of the population was a victim of violence in the last 12 months. However, no statistical relationship existed between that phenomenon and the consistent or inconsistent use of condoms (Clements-Nolle et al., 2008). In Poteat’s investigation, 17.2% of TW were tortured, 33.2% were attacked, and 24.4% felt scared to walk in public places (Poteat et al., 2017). Mburu et al. (2019) presented an association between the TW physically abused (23.6%) and higher use of amphetamine-type stimulants. Raiford et al. (2016) presented results of physical and verbal abuse; all the TW reported verbal victimization and 87% physical victimization at some point in their life. However, the authors reported a decrease of 30% and 10%, respectively, recently (Raiford et al., 2016).

##### Frequent alcohol use

Regarding the frequent use of alcohol, we found twenty-eight studies that presented a variation from 7.7% (Mimiaga et al., 2019) to 67.8% (Alvarado et al., 2020) (Supplementary Table 2). Avila et al. (2017) also reported that 32.3% of TW said that alcohol facilitates sex work, and 20% expressed a relation between condom use and alcohol (Avila et al., 2017). Denson et al. (2017) showed that 37.3% of TW had their last sexual intercourse with a man under the effects of alcohol or drugs (Denson et al., 2017). Ferreira et al. (2016) also showed high use of alcohol among the TW (60%). Mburu et al. (2019) found the correlation that the more alcohol they drank, the higher their amphetamine-type stimulants consumption. Similarly, alcohol use is related to unprotected sex with the primary male partner (Mburu et al., 2019).

The study by Shrestha et al. (2020) reported that 17.5% of TW consumed alcohol in the last 30 days, and Poteat et al. (2017) showed that TW consumed a mean of 2.91 alcoholic drinks in one sitting.

##### Illicit drug use

Twenty-three studies reported percentage data of illicit drug use by TW (Supplementary Table 2). The use of drugs was frequent among the study participants, according to Operario et al. (2008). Ferreira et al. (2016) and Avila et al. (2017) studies reported that 43.9% and 33.2% (respectively) of their TW participants used illegal drugs. Ferreira et al. (2016) also found that 52.3% of those who used illegal drugs declared that they had an influence on sex work, and 33.3% declared an influence on condom use. Wickersham et al. (2017) also found the use of illegal drugs during sex work, 33.2% of TW used drugs in the last 30 days; however, that data was almost equal to the cisgender population (33.8%) (Wickersham et al., 2017). Clements-Nolle’s study analyzes the specific weight of several kinds of illegal drugs in the use of condoms (Clements-Nolle et al., 2008), and Denson et al. (2017), describe the use of drugs by type of administration (injected/no injected), and the relation of those behaviors with others, as having sex under the effects of drugs or alcohol. The study by Mburu et al. (2019) focused on amphetamine-type stimulants and their relation to health behaviors, depressive symptoms, gender-related discrimination, violent experiences, and childhood abuse.

Finally, Poteat et al. (2017) presented the use of drugs to other risk behaviors, such as sharing needles. 40% of TW ever shared needles, but only 1.1% did it in the last year. Not injection drug use was 17% among the same population (Poteat et al., 2017).

##### Financial instability

Twenty-four studies showed percentage data of financial instability self-reported by TW (Supplementary table 2). Raiford et al. (2016) explored multiple variables related to financial instability, such as age, education, and income, and the impact of those on risky behaviors (Raiford et al., 2016). Clements-Nolle et al. (2008) show an association between socioeconomic factors and the use of condoms: higher educational level and higher income are related to consistent use. Operario et al. (2008) found a relationship between unemployment and unprotected sex. Denson’s evidence showed that most of the population was unemployed or disabled for work (60%), and only 15% had full-time jobs. Relatively, 58.1% had a household income lower than 10,000 USD (Denson et al., 2017). Mburu compares the labor environment and the use of drugs, evidencing that 42% of TW had problems getting a job due to their identity, and 24.3% have lost a job for the same reason (Mburu et al., 2019).

##### Depression

Nine studies showed percentage data of depression self-reported by TW (Supplementary Table 2). Poteat et al., (2017) compared the depression experienced by TW about other populations, using a scale from 0-60. They found that TW reported a mean of 16.8, lower than the cisgender woman (Poteat et al., 2017). On the other hand, other studies (Clements-Nolle et al., 2008; Operario et al., 2008) showed a prevalence of depression in TW of 58.6%, and a relation with unprotected sex on 66.7% of them. Shrestha et al. (2020) identified that 56% of the TW participants in their study experimented with depression, and there was a slight tendency to reject the use of self-testing HIV devices.

##### Unstable housing

Ten studies showed percentage data of unstable housing self-reported by TW (Supplementary Table 2). The data relating to housing stability differed in each investigation, ranging from 11% (Wickersham et al., 2017) to 49% (Clements-Nolle et al., 2008). Moreover, the study by Denson et al. (2017) showed that 46.3% of TW were homeless at some point in the last 12 months. Avila et al. (2017) reported a percentage of 29.6%, Operario et al. (2008) found 29.8%, almost one third of TW sex workers, lived in precarious places, Raiford et al. (2016) found 21%, and Ferreira et al. (2016) deduced closer to 15.2%.

##### Completed high school

Forty-one studies showed percentage data of completed high school studies self-reported by TW (Supplementary Table 2), presenting a variation from 9.4% (Magno et al., 2018) to 79.7% (Alvarado et al., 2020). Shrestha et al. (2020) described that 69.3% of their study population completed high school. Ferreira et al. (2016) found 69.7% of TW completed high school, Clements-Nolle et al. (2008) found data closer to 65%, and Denson et al. (2017) reported 65.7% completed high school. Furthermore, Avila et al. (2017) found an association between completed high school (54.9%) and a lower prevalence of syphilis. Similar data related to the educational level was found by Wickersham et al. (2017), with 54.4%, and Raiford et al. (2016), with 54%.

##### Stigma and discrimination

Fifteen studies reported stigma and discrimination data self-reported by TW. Data from this variable was often divided into different areas such as family, friends, and school communities (Data not shown in supplementary Table 2). Mburu et al. (2019) evidenced a high level of support (89.8%) from their co-workers/classmates, but 24.2% of them dropped out of school, 18.1% had housing issues, and 10% were arrested because of their identity. Raiford et al. (2016) identified the stigma perceived by the TW on a scale from 10-40, with a mean of 23.3. Likewise, Poteat et al. (2017) reported several stigma events suffered by TW, including being afraid to seek medical assistance (26.7%). However, only 2.2% reported having been denied medical care. The authors also analyzed discriminatory behaviors in familiar contexts finding that 18.3% were excluded from gatherings, 35.1% suffered gossip or discriminatory remarks, and friendship rejection of 30% due to sexual orientation.

## Discussion

The social factors have a proven relationship with condoms and HIV among TW. The syndemic factors, their complexity of layers, the different weights of each one, and how some of them reinforce others (i. e. the stigma and their impact on the school drop-off and subsequent low-income, homeless, and drug use, among others) deserve a specific analysis to understand, design, and develop strategies to prevent the spread of HIV (Mburu et al., 2019; Raiford et al., 2016). The population of the included studies represents different cultures and political contexts, finding that some factors (such as financial instability or childhood sex abuse) constantly threaten the well-being of TW (Clements-Nolle et al., 2008; Denson et al., 2017; Shrestha et al., 2020).

In this work, the referred authors highlight the relevance of the syndemic factors as an epistemological, theoretical, and empirical perspective to describe and analyze “how social and health conditions interact in a significant way through social, biological, and psychological pathways, and are driven by social and political forces” (Mendenhall, 2020). This crossroads of conditions and constraints configures a nucleus of problems associated with health, illness, and attentional processes. Thus, the use or not of a condom by trans women is a phenomenon that should not be read from a single and reductionist perspective, but from the multidimensionality that the mentioned experience goes through.

This review shows, about multiple adversities faced by TW, that the decision to have or not have unprotected sex loses relevance, not necessarily as a rational choice, but due to the imposition of conditions that reduce the margin of action and have different relational logics and even overwhelming with these women. The syndemic perspective explains why the risks associated with unprotected sexual intercourse take a backseat for TW, as they represent a population group with multiple disadvantages. HIV and the conditions in which it occurs to respond to individual, collective, social, economic, and cultural phenomena. Consequently, it is from this broad framework that it must be addressed as a public health problem.

According to some studies reviewed, phenomena such as childhood sexual abuse and violence victimization may impact the TW before “condom use” is the measured risky behavior. Moreover, previous studies (Li et al., 2014; Willie et al., 2018; Willie et al., 2022) have reported a relationship between women that suffer violence and an increased risk for HIV acquisition; however, we believe this possible relation in TW requires further investigation.

## Limitations

It is necessary to explore more about the specific weights of each phenomenon and, at the same time, evaluate the impact of the current wave of policies (such as the gender policies or anti-discrimination initiatives at schools) on the well-being of TW.

## Conclusions

Regarding HIV’s effect on TW, there is an opportunity to know and do more about sexual health behaviors. The use of condoms was reported in less than 60% of the studies, but as relevant as that is, the fact that there are other phenomena faced by TW that increase HIV contraction risks. The syndemic perspective opens the opportunity to understand and impact the health of TW before the risky behavior and protect them during their lifetime (work life, health services, prison) by creating specific policies. As a public health problem, the risk is not created by the no-use of condoms, but rather has multiple determinants and requires further research and action to protect TW.

Assessing syndemic factors provides insight into the complex relationship between TW’s real-life context and their sexual, mental, and physical health in different ways. First, it helps to question the stigma that directly associates TW with HIV and the repercussions that this brings to their mental health. Secondly, it favors identifying the factors most related to the decisions of condom use in the sexual practices of TW. This is key since a large majority of articles have investigated the trans population together with the MSM or LGB population, which does not allow differentiating between the specific needs of each group. Thirdly, we consider that the analysis of syndemic factors is necessary to generate health intervention and prevention programs aimed at reducing HIV that are more contextualized to the realities of TW.

## Supporting information

PRISMA ScR checklist

## Data Availability

The authors confirm that the data supporting the findings of this study are available within the article and its supplementary materials.

