## Supplementary material 1 for "Characterization and contextualization of transgender women population and condom use in the HIV syndemic framework: Scoping review"

Supplementary material 1. Characteristics of all included studies.

| **Author** | **Year** | **Country** | **Study design** | **Total sample size** | **n_TW** | **Ethnicity (%)** | **Age (mean, years)** | **Sex worker (%)** |
| --- | --- | --- | --- | --- | --- | --- | --- | --- |
| Alvarado et al. | 2020 | Colombia | Cross-sectional | 812 | 59 | Afrocolombian or indigenous 30.5 | **-** | **-** |
| Avila et al. | 2017 | Argentina | Cross-sectional | 273 | 273 | - | - | 100.0 |
| Bao et al. | 2016 | Vietnam | Cross-sectional | 204 | 204 | Kinh 93.1  Other 6.9 | 18-41 | - |
| Bhatta et al. | 2014 | Nepal | Cross-sectional | 232 | 232 | - | 25.0 | - |
| Bowers et al. | 2012 | USA | Prospective | 912 | 255 | White 12.9 African American 18.4 Latino 50.6 Other 18 | 31.9 | - |
| Budhwani et al. | 2017 | Dominican Republic | Prospective | 78 | 78 | - | 23.0 | 100.0 |
| Cai et al. | 2016 | China | Cross-sectional | 220 | 183 | - | - | 100.0 |
| Chakrapani et al. | 2018 | India | Cross-sectional | 300 | 300 | - | 29.6 | 70.7 |
| Clements-Nolle et al. | 2008 | USA | Cross-sectional | 190 | 190 | African American 28.0 Latino 31.0 White 22.0 Other 18.0 | 31.5 | 100.0 |
| Colby et al. | 2016 | Vietnam | Cross-sectional | 205 | 205 | Kinh 93  Chinese 5  Khmer 1 | 25.0 | - |
| Degtyar et al. | 2018 | Peru | Ethnographic mapping | 376 | 181 | - | 25.0 | 100.0 |
| Denson et al. | 2017 | USA | Cross-sectional | 241 | 241 | African American 61.2 Latino 38.8 | 15-40 | - |
| Drückler et al. | 2020 | the Netherlands | Retrospective | 164 | 15 | Nonwestern 73.3 Western: 26.7 | 39.0 | 100.0 |
| Ferreira Jr et al. | 2016 | Brazil | Cross-sectional | 124 | 66 | White: 40,9  Other: 59.1 | 18-50 | - |
| Forbes et al. | 2016 | USA | Prospective | 105 | 59 | African American 8.5  White 5.1 Latina 49.2 Asian Pacific Islander 16.9 Other 13.6 Missing 6.8 | 20.7 | 37.8 |
| Gama et al. | 2018 | Portugal | Cross-sectional | 125 | 125 | - | 32.0 | 100.0 |
| Hearld et al. | 2019 | Dominican Republic | Retrospective | 291 | 291 | - | 26.0 | 37.1 |
| Kattari et al. | 2019 | USA | Retrospective | 4834 | 42 | White 53.8  Other 46.2 | 16.2 | - |
| Khalid & Martin | 2018 | Pakistan | Retrospective | 2326 | 711 | - | 28.0 | 100.0 |
| Khan et al. | 2008 | Pakistan | Prospective | 409 | 409 | - | 24.0 | 100.0 |
| Logie et al. | 2020 | Jamaica | Cross-sectional | 340 | 101 | - | 24.6 | 100.0 |
| Logie et al. | 2019 | Jamaica | Cross-sectional | 911 | 137 | - | 24.2 | - |
| Long et al. | 2019 | Peru | Cross-sectional | 3336 | 142 | - | 29.6 | 65.2 |
| Long et al. | 2019 | Peru | Retrospective | 310 | 310 | - | 28.9 | - |
| Magno et al. | 2018 | Brazil | Prospective | 127 | 127 | White 18.9 African American 29.9 Brown 51.2 | - | 87.4 |
| Maliya, Irwan, Samsul, Zakiah, Faridah | 2018 | Malaysia | Cross-sectional | 54 | 54 | - | 39.4 | 25.0 |
| Mburu et al. | 2019 | Cambodia | Retrospective | 1375 | 1375 | - | 25.8 | - |
| Mimiaga et al. | 2019 | USA | Randomized controlled trial | 233 | 233 | White 25.8  Other 74.2 | 23.4 | - |
| Moayedi et al. | 2019 | Iran | Prospective | 104 | 104 | - | 27.9 | - |
| Moriarty et al. | 2019 | Peru | Retrospective | 120 | 120 | - | 29.5 | - |
| Murphy et al. | 2020 | Peru | Retrospective | 389 | 389 | - | 26.0 | 62.2 |
| Nemoto et al. | 2011 | Thailand | Cross-sectional | 112 | 112 | Thai 96.4 Other 3.6 | 25.0 | 100.0 |
| Operario et al. | 2011 | USA | Cross-sectional | 174 | 174 | African American 24.0 Hispanic 23.0 Asian 17.0 White 12.0 Mixed-race, other 23.0 | 37.8 | - |
| Parsons et al. | 2018 | USA | Cross-sectional | 212 | 212 | African American, 31.6  Latino 3.5 White 24.5 Mixed-race, other 10.4 | 34.3 | 100.0 |
| Poteat et al. | 2019 | Thailand and Brazil | Cross-sectional | 353 | 37 | - | 19-54 | 10.8 |
| Raiford et al. | 2016 | USA | Retrospective | 63 | 63 | Hispanic 46.0 African American, non-Hispanic 35.0 White, non-Hispanic 6.0 Other 13.0 | 38.1 | - |
| Rana et al. | 2016 | Bangladesh | Cross-sectional | 889 | 889 | - | > 15 | 73.3 |
| Reback et al. | 2018 | USA | Retrospective | 271 | 271 | Latino 42.1  African American 30.3  Other 27.7 | 18-40 | 100.0 |
| Richter et al. | 2013 | South Africa | Cross-sectional | 1799 | 59 | - | 28.7 | 100.0 |
| Satcher et al. | 2017 | Peru | Retrospective | 138 | 138 | Latino 100.0 | 27.0 | - |
| Shan et al. | 2018 | China | Cross-sectional | 498 | 498 | - | 30.0 | - |
| Shrestha et al. | 2020 | Malaysia | Cross-sectional | 361 | 361 | - | 35.3 | 100.0 |
| Stahlman et al. | 2016 | Ivory Coast, Togo and Burkina Faso. | Cross-sectional | 2456 | 453 | - | 22.5 | 48.1 |
| Storm et al. | 2020 | Nepal | Retrospective | 340 | 173 | - | 31.8 | 100.0 |
| Turner et al. | 2017 | USA | Cross-sectional | 263 | 263 | White 39.9 Latino 31.6 African American 12.2 Other 16.4 | 21.2 | - |
| Weissman et al. | 2016 | Cambodia | Cross-sectional | 891 | 891 | - | 23.0 | 3.7 |
| Wickersham et al. | 2017 | Malaysia | Cross-sectional | 492 | 193 | Malay 58.5  Indian 29.5  Indonesian 10.4  Chinese 0.5  Other 1.0 | 34.3 | 100.0 |
