## Supplementary material 2 for "Characterization and contextualization of transgender women population and condom use in the HIV syndemic framework: Scoping review"

Supplementary material 2. Included studies that reported condom use data.

| **Author** | **Time window/situation** | **Frequency** | **General Condom Use % (n)** | **Condom use with stable partner % (n)** | **Condom use with casual partners % (n)** | **Condom use with clients % (n)** |
| --- | --- | --- | --- | --- | --- | --- |
| Alvarado et al. | - | Yes or no | 38.9 (59) | 34.4 (59) | 60.5 (59) | 65.2 (59) |
| Avila et al. | Last 6 months | regularly (always use condom) and irregularly (sometimes or never use condom). | - | 34.6 (78) | - | 64.6 (268) |
| Bao et al. | Last month | Never  Usually/sometimes  Always | - | - | Test HIV positive 45.9 (74) Test HIV negative 24.3 (37) | Test HIV positive 58.3 (48) Test HIV negative 21.7 (23) |
| Budhwani et al. | The last time you had sex, you used a condom | Yes or no | - | 67.8% (42) | 92.96% (66) | 91.78 (67) |
| Clements-Nolle et al. | Last 6 months | Five-point Likert scale (always, almost always, sometimes, rarely, never) and further dichotomized into consistent (always) and inconsistent (less than always) categories | 80.5 (190) | - | - | - |
| Colby et al. | Last month | 4-item Likert scale of never, sometimes, usually, or always. The questionnaire did not differentiate between insertive or receptive sex. Based on a post hoc review of the data, consistent condom use was defined as habitual or always using condoms; condomless sex (CLS) was defined as never or sometimes reporting condom use. |  | 25 (97) | 42 (102) | 49 (67) |
| Degtyar et al. | Last 3 months |  | 69.0 | - | 48.0 (56) | 69.0 (179) |
| Denson et al. | Last 12 months | Yes or no | 51.4 (212) | - | - | - |
| Drückler et al. | Last 6 months |  | 66.7 (15) | - | - | 80.0 (15) |
| Ferreira Jr et al. | - | Yes or no | - | 40.9 | 77.3 | - |
| Hearld et al. | Last month | Always, Sometimes or never | 86.22 | - | - | - |
| Kattari et al. | Last time | Yes or no | Did not use condom 59.52 | - | - | - |
| Khalid & Martin | Last month | Always, sometimes or never | 45.7 | - | - | - |
| Logie et al. | Last month | Consistent or inconsistent | 70.2 | 71.6 | 77.8 | 78.6 |
| Long et al. | Last 3 months | - | Insertive anal sex without a condom 50.8  Receptive anal sex without a condom 82.8 | Sex without a condom  52.6 | Sex without a condom 40.0 | Sex without a condom  0.0 |
| Long et al. | Last time | - | 49.0 | - | - | 74.9 (188) |
| Magno et al. | - | Always, most of the times, rarely or never | - | 71.6 (116) | 93.7 (127) | 99.1 (111) |
| Maliya et al. | Last month | Every time, often, sometimes, rarely, never | - | 29.4 (15) | 29.4 (15) | 33.3 (17) |
| Mburu et al. |  | Always or Not always | - | - |  | 58.4 |
| Moayedi et al. | Last time | Always or most of the time | 39,7% (29) | 39.7 | 34.6 | 53.3 |
| Poteat et al. 2017 | Last time | - | - | 70.1 | 76.7 | - |
| Rana et al. | Last time | - | 58.0 (516) | - | Condom used in last non-transactional anal intercourse in the last month  40.6 (279) | Condom used in last anal intercourse with new clients in the last week  64.4 (310)  Constantly used a condom during anal intercourse with new clients in the last week  35.1 (169)  Condom used in last anal intercourse with regular clients in the last week  65.1 (339)  Consistently used a condom during anal intercourse with regular clients in the past week  34.0 (177) |
| Shan et al. | Last 3 months | Always | - | 18.50 | 29.10 | 14.0 |
| Storm et al. | Last time | Yes or no | 55.5 | - |  |  |
| Weissman et al. | Last time | Yes or no | 83.9 (763) | - | 44.9 |  |
| Wickersham et al. | - | - | 89.0 (168) | - | - | - |
