## Supplementary material 3 for "Characterization and contextualization of transgender women population and condom use in the HIV syndemic framework: Scoping review"

Supplementary material 3. Syndemic factors of included studies

| **Author** | **Syndemic factor % (n)** | | | | | | | | | |
| --- | --- | --- | --- | --- | --- | --- | --- | --- | --- | --- |
|  | **History of incarceration** | **Childhood sexual abuse** | **Forced sex** | **Violence victimization** | **Frequent alcohol use** | **Illicit drug use** | **Financial instability** | **Depression** | **Unstable housing** | **Completed high school** |
| Alvarado et al. | 0.0 (59) | 52.4 (59) | 6.9 (59) | **-** | 67.8 (59) | 59.3 (59) | 61 (59) | **-** | **-** | 79.7 (59) |
| Avila et al. | 67.9 (268) | 5.9 (221) | 25.7 (226) | Being beaten and have HIV test  45.3 (265) | 62.7 (271) | 33.2 (265) | - | - | 29.6 (270) | 54.9 |
| Bao et al. | - | - | - | - | HIV positive 72.2 (97) HIV negative 66.6 (69) | HIV positive  36.7 (120) HIV negative  28.9 (83) | HIV positive 33.1 (121) HIV negative 24.1 (83) | - | - | HIV positive  52.5 (120) HIV negative  41.0 (83) |
| Bhatta et al. | - | - | - | - | 68.5 | - | 25.9 | - | - | 57.3 |
| Bowers et al. | - | - | - | - | 11.7 (74) | 19.9 (125) | - | - | 58.0 (148) | 54.3 (138) |
| Budhwani et al. | - | - | - | 42.3 | - | - | - | - | - | Secundary  37.2 College or University  35.9 |
| Cai et al. | - | - | - | - | - | - | 25.9 | - | - | Primary or below  11.9 Junior secondary  35.9 Senior secondary  41.4 Tertiary  10.9 |
| Chakrapani  et al. | - | - | - | 83.7 | 37.3 | - | Average monthly income 8.071,3 INR  [In USD, mean=134, SD=104] | 35.7 | - | Non-formal education  14.3 Up to secondary education  63.3  Above secondary education  22.3 |
| Clements-Nolle et al. | 43.0 | - | 63.0 | IPV 19.0 (36) | - | Cocaine  28.0 Crack-cocaine  31.0 Marihuana  69.0 Methamphetamines 43.0 Injected drug  24.0 | 21.0 | 58.0 | 49.0 | 65.0 |
| Colby et al. | - | - | 23.0 | - | 91.0 | - | 30.0 | 45.0 | - | Primary and below  25.0  Middle school  28.0 High School  35.0 University  13.0 |
| Degtyar et al. | - | - | - | - | - | - | - | - | - | 36.0 |
| Denson et al. | 23.8 | - | 15.5 | - | 65.3 | 38.0 | 58.1 | - | 46.0 | 65.7 |
| Drückler et al. | - | - | - | - | 86.7 | 60.0 |  | - | - | - |
| Ferreira Jr et al. | 12.1 | - | - | - | 60.6 | 43.9 | - | - | 20.0 | 0-4 years  3.0  5-8 years  27.3  9-11 years  37.9  12 or more years  31.8 |
| Forbes et al. | - | - | - | - | - | - | - | - | - | 27.1 |
| Gamarel et al. |  |  |  |  |  |  |  |  |  |  |
| Hearld et al. | - | - | - | Verbal abuse 47.9  Physical abuse 14.1 | 48.1 | - | 44.8 | - | - | - |
| Kattari et al. | - | - | - | - | 19.7 | 19.7 | - | - | - |  |
| Khalid & Martin | - | - | - | - | - | - | - | - | - | 28.0 |
| Khan et al. | - | - | - | 40.0 | 20.0 | 43.0 | - | - | - | - |
| Logie et al. | - | - | - | - | 28.7 | - | - | - | - | 78.2 |
| Logie et al. | - | 34.3 | - | 26.5 | - | - | - | - | 53.0 | 80.3 |
| Long et al. | - | - | - | - | 31.0 | - | - | - | - | 39.4 |
| Long et al. | - | - | - | - | 33.2 | - | - | - | - | 32.9 |
| Magno et al. | - | - | - | Any violence from Police  47.2 Physical Violence  59.1 | - | - | 35.4 | - | - | 9.4 |
| Maliya, Irwan, Samsul, Zakiah, Faridah | - | - | - | - | - | - | 7.4 | - | - | 63.5 |
| Mburu et al. | 10.5 | 32.5 | 39.1 | 23.6 | 51.3 | 10.4 | 63.7 | 66.8 | - | 36.6 |
| Mimiaga et al. | - | 10.0 (140) |  | IPV  41.7  Victimization by transgender people  74.9 | 7.7 | 15.9 | - | 42.1 | - | High school/equivalent or less  45.1  Some college  18.5  College  3.4  Some graduate school or more  2.6  Current student  30.5 |
| Moayedi et al. | - | 24.7 (73) | - | - | 36.5 | 8.7 | - | - | - | 79.8 |
| Moriarty et al. | - | - | - | - | 64.2 | 60.0 | 40.8 | - | - | 24.2 |
| Murphy et al. | - | - | 2.30 | 17.5 | - | - | 45.3 (243) | - | - | 80.7 (343) |
| Nemoto et al. | - | 43.80 | - | 43.8 | 99.1 | 97.2 | 3.2 | - | - | 61.6 |
| Operario et al. | 52.3 | - | - | - | 58.0 | 62.6 | 62.4 | 58.6 | 29.8 | 74.3 |
| Parsons et al. | - | 41.5 | - | 65.1 | - | 33.0 | 80.2 | 41.5 | - | 58.4 |
| Poteat et al. 2017 | Been arrested  7.9 (72) Been in jail 5.2 (48) | - | 27.6 | Physically beaten/attacked  33.2 (311) Tortured  17.2 (74) | Mean number of drinks in one sitting  2.91 | Inject drugs in the last 12 months  1.1 (5)  Take any medication that was not injected and not prescribed in the last 12 months  17.9 (166) | - | 57.3 | - | - |
| Raiford et al. | 76.0 | - | 100.0 | Verbal  100  Physical  87 | - | - | 54 | - | 21.0 | 54.0 |
| Rana et al. | - | - | - | - | - | 14.3 | Average income of BTk 7000 in the last month  51.1 | - | - | 57.4 |
| Reback et al. | - | - | - | 56.8 | 40.9 | - | 83.7 | - | - | 63.5 |
| Richter et al. | - | - | - | - | 66.5 | - | - | - | - | Complete High School  33.3 (18) Received tertiary education  7.4 (4) |
| Satcher et al. | - | - | - | - | 65.7 | - | - | - | - | 66.1 |
| Shan et al. | - | - | - | - | - | 40.2 | - | - | - | ≤ 9 years  13.9 (69)  10–12 years  29.1 (145)  > 13 years  57.0 (284) |
| Shrestha et al. | 36.0 | 41.3 | - | - | 17.5 | 10.2 | 13.9 | 56.0 | - | 69.3 |
| Stahlman et al. | - | - | 21.9 | 25.3 | - | - | 11.3 | - | - | Primary school or lower  16.1 (72)  Secondary/high school  58.7 (263)  More than high school  25.2 (113) |
| Storm et al. | - | - | 18.5 | 11.6 | - | - | 19.7 | - | 26.0 | Ever attended school  75.7 |
| Turner et al. 2017 | - | - | - | 61.7 | - | - | - | - | - | 54.0 |
| Weissman et al. | - | - | 30.3 (880) | 30.3 (880) | 13.5 (873) | 9.3 (870) | 50.2 (447) | - | - | Never attended school  3.3 (29)  Primary level  14.5 (126)  Lower secondary  25.5 (222)  Secondary  31.7 (276)  University & higher  24.9 (217) |
| Wickersham et al. | 50.3 (97) | - | - | - | - | 33.2 (64) | - | - | 11.4 | 54.4 |

IPV: Intimate partner violence
